# Diagnosis of Acute Respiratory Syndrome Coronavirus 2 Infection by Detection of Nucleocapsid Protein

**DOI:** 10.1101/2020.03.07.20032524

**Authors:** Bo Diao, Kun Wen, Jian Chen, Yueping Liu, Zilin Yuan, Chao Han, Jiahui Chen, Yuxian Pan, Li Chen, Yunjie Dan, Jing Wang, Yongwen Chen, Guohong Deng, Hongwei Zhou, Yuzhang Wu

## Abstract

**BACKGROUND:** Nucleic acid test and antibody assay have been employed in the diagnosis for SARS-CoV-2 infection, but the use of viral antigen for diagnosis has not been successfully developed. Theoretically, viral antigen is the specific marker of the virus and precedes antibody appearance within the infected population. There is a clear need of detection of viral antigen for rapid and early diagnosis.

**METHODS:** We included a cohort of 239 participants with suspected SARS-CoV-2 infection from 7 centers for the study. We measured nucleocapsid protein in nasopharyngeal swab samples in parallel with the nucleic acid test. Nucleic acid test was taken as the reference standard, and statistical evaluation was taken in blind. We detected nucleocapsid protein in 20 urine samples in another center, employing nasopharyngeal swab nucleic acid test as reference standard.

**RESULTS:** We developed a fluorescence immunochromatographic assay for detecting nucleocapsid protein of SARS-CoV-2 in nasopharyngeal swab sample and urine within 10 minutes. 100% of nucleocapsid protein positive and negative participants accord with nucleic acid test for same samples. Further, earliest participant after 3 days of fever can be identified by the method. In an additional preliminary study, we detected nucleocapsid protein in urine in 73.6% of diagnosed COVID-19 patients.

**CONCLUSIONS:** Those findings indicate that nucleocapsid protein assay is an accurate, rapid, early and simple method for diagnosis of COVID-19. Appearance of nucleocapsid protein in urine coincides our finding of the SARS-CoV-2 invading kidney and might be of diagnostic value.

## Introduction

Acute respiratory syndrome coronavirus 2 (SARS-CoV-2) was named by the world health organization in 2019, causing COVID-19[1]. By March 6, 2020, 80,714 cases were laboratory confirmed and 3045 were death in China based on CCDC reported. SARS-CoV-2 is an enveloped RNA virus widely distributed in humans, other mammals and birds that cause respiratory, intestinal, liver and neurological diseases [2, 3]. Six coronaviruses are known to cause disease in humans. SARS-CoV-2 is a novel coronavirus that has never been found in humans. Common symptoms in people infected with SARS-CoV-2 virus include respiratory symptoms, fever, cough, and in more severe cases, infection can lead to pneumonia and severe acute respiratory syndrome [4,5]. The known routes of coronavirus disease-19(COVID-19) transmission are respiratory droplets and contact transmission, while aerosol and fecal-oral transmission require further clarification [6].

For SARS-CoV-2 infection, early diagnosis is particularly important, not only to prolong the survival of patients, but also to ensure the safety of the population [7]. In clinical practice, the detection methods have been rapidly updated due to the deepening understanding of COVID-19. Nucleic acid testing, chest CT, confirmation of epidemiological history and clinical manifestations are important bases for the diagnosis of novel coronavirus pneumonia [7-10]. However, nucleic acid testing has operator restrictions, time-consuming, easy to pollution and the high cost. In comparison, the coronavirus antigen detection method has the advantages of rapid, convenient and the SARS-CoV antigen can be detected up to 1 day before appearance of clinical symptoms [11].

In clinical practice, nucleic acid test and antibody assay have been employed in the diagnostic for suspected acute respiratory syndrome coronavirus 2 (SARS-CoV-2) infections, but the use of SARS-CoV-2 antigen for diagnosis has not been successfully developed. Theoretically, viral antigen is the specific marker of the virus and precedes antibody appearance within the infected people. Therefore, detection of viral antigen can play a rapid screening effect and achieve the purpose of early diagnosis. Here, we report a fluorescence immunochromatographic assay for detecting nucleocapsid protein of SARS-CoV-2 in nasopharyngeal swab samples and urine within 10 minutes, and evaluated its significance in diagnosis of COVID-19.

## Methods

### Study design

A total of 239 participants with suspected SARS-CoV-2 infection from 7 centers, including General Hospital of Central Theater Command, Wuhan N0.7 People’s Hospital, Wuhan Pulmonary Hospital,Hubei Maternal and Child Hospital,Taikang Hospital,Hanyang Hospital and Wuguo Hospital, were employed in this clinical trial for diagnosis. Nucleic acid test was taken as golden standard. We measured nucleocapsid protein in nasopharyngeal swab samples in parallel with the nucleic acid test. Statistical evaluation was taken in blind based a four-grid table. We detected nucleocapsid protein in 20 urine samples in Southwest Hospital in Chongqing, employing parallel nasopharyngeal swab nucleic acid test as reference standard for the same day. This study was approved by the ethics commission of participated hospitals.

### Sample selection

Inclusion criteria: nasopharyngeal swab samples and urine from suspected cases of COVID-19. Exclusion criteria: contaminated samples; duplicate samples; unclear samples; samples with missing information from the original records of clinical trials; samples conditions do not meet the program requirements.

### Participants

Medical records of 239 participants in Wuhan and 20 participants in Chongqing were collected and retrospectively analyzed.

### Nucleic acid test of SARS-CoV-2

Nasopharyngeal swab specimen were collected from all participants. Extraction of nucleic acids from those samples performed with a High Pure Viral Nucleic Acid Kit, as described by the manufacturer (Da An Gene Co., Ltd). Extracted nucleic acid samples were tested for viruses and bacteria by polymerase chain reaction (PCR), using the ABI Prism 7500 and the Light Cycler 480 real-time PCR system, in accordance with manufacturer instructions. A real-time reverse transcription PCR (RTPCR) assay was used to detect viral RNA by targeting SARS-CoV-2 ORF1ab and N gene region of SARS-CoV-2.

### Nucleocapsid protein detection by fluorescence immunochromatographic assay

The test device, which is developed based on the principle of fluorescence Immunochromatographic assay, is composed of a sample pad, a conjugate pad, a nitrocellulose membrane, an absorbent pad, and a supporting plastic cassette (Fig. 1A). The goat anti-rabbit IgG antibodies (2mg/mL) and mouse anti-nucleocapsid protein of SARS-CoV-2 monoclonal antibody (M1,1mg/ml) were dotted on the nitrocellulose membrane at a volume of 0.4 μL to form the control line and test line, respectively. The conjugates of carboxylate-modified polystyrene Europium (III) chelate micro particles with anti-nucleocapsid protein of SARS-CoV-2 monoclonal antibody M4 or rabbit IgG (100 μg/mL) were spotted on the conjugation pad at a volume of 1.2 μL. The fully assembled immunochromatographic strip was then sealed into the plastic cassette (Fig. 1B) with a sample well and this test device was stored in a dry environment at room temperature and ready for further experiment. With the help of the capillarity of the absorbent pad, sample buffer containing SARS-CoV-2 nucleocapsid protein could be combined by III-conjugated M4 antibody and captured by M1 antibody at the test line, while III-conjugated rabbit IgG could be captured on the control line, resulting in a fluorescent band on the test and control lines, respectively. The fluorescent results will then be read by the immunofluorescence analyzer. Cut-off value was determined by testing 100 nasal swab samples of healthy people and calculated as the mean value of the fluorescence signal plus 5 SD.

### Nucleocapsid protein detection procedure

Nasopharyngeal swab samples or urine were diluted and mixed well with 500μl saline solution, 100μl of which was then added to the sample well of the test card. After a ten-minute reaction, immediately inserted the card into the immunofluorescence analyzer (Fig. 1D) and the instrument could automatically determine the positive or negative result by comparing the detection value with the reference cut-off value setting within the internal parameter of the kit’s ID chip (Fig. 1C).

**Fig. 1.**
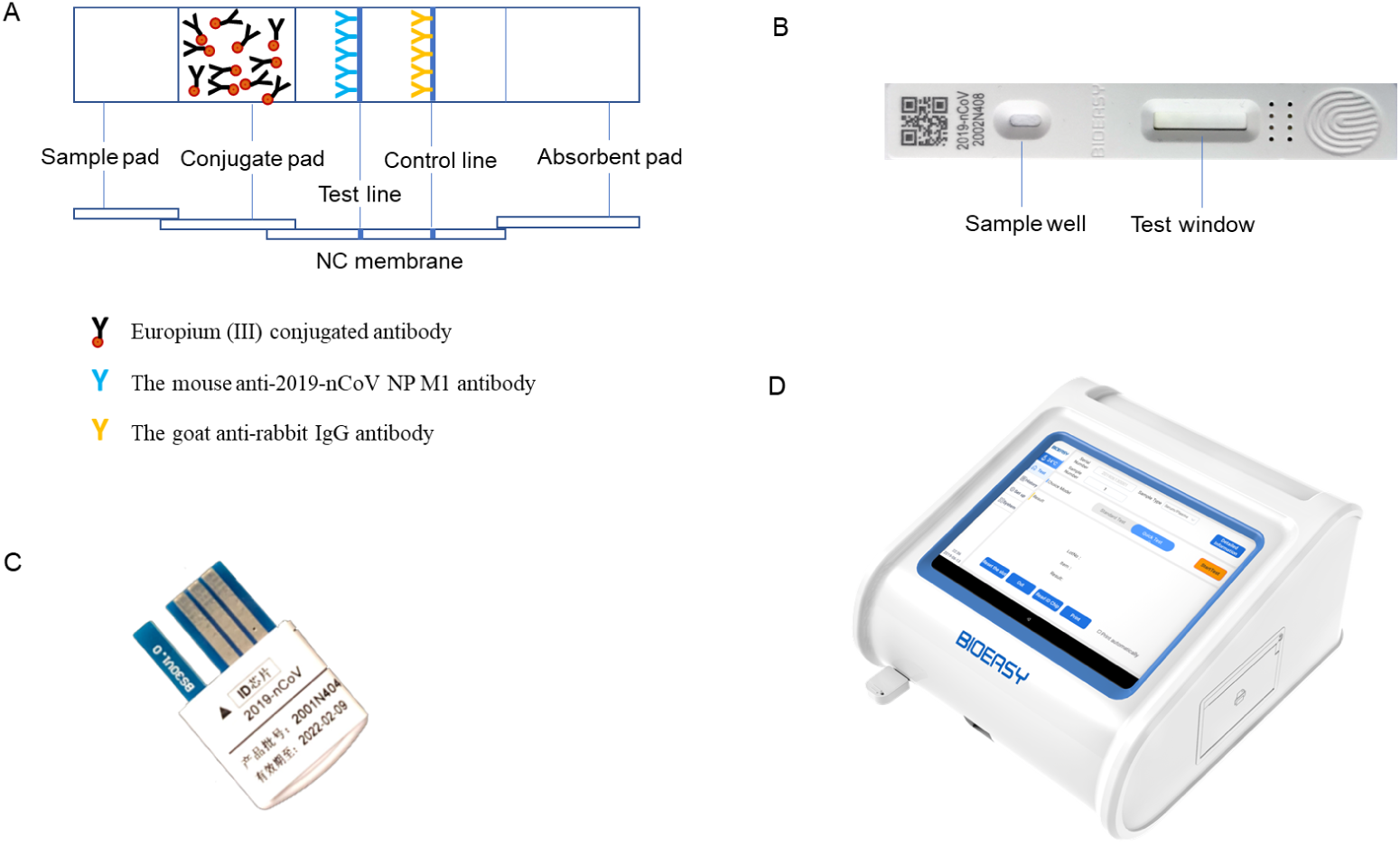
**(A)** Description of the fluorescence immunochromatographic strip. **(B)** Test device. **(C)** ID chip. **(D)** Immunofluorescence analyzer.

### Data analysis

Statistical analyses were performed blindly with the statistical analysis system software SPSS 19.0.

### Role of funding source

This research was supported by grants from National Key R&D Program of China (2016YFA0502204); Chongqing Health Commission COVID-19 Project (2020ZX01). The corresponding authors were responsible for all aspects of the study to ensure that issues related to the accuracy or integrity of any part of the work were properly investigated and resolved. The final version was approved by all authors.

## Results

### 1. Clinical features and COVID-19 nucleic acid detection

Our study enrolled 239 participants based on diagnostic criteria for suspected SARS-CoV-2 infection. All participants underwent 3 nucleic acid tests, and the results of each nucleic acid test were verified by 2 COVID-19 nucleic acid test kits. As shown in Fig 2A, among the 239 suspected COVID-19 patients, COVID-19 nucleic acid cycle threshold (CT value) of 208 patients≤40, while 31 patients>40, indicating 208 patients tested positive for nucleic acid and 31 patients tested negative for nucleic acid, As shown in Fig 2B, among the 239 suspected COVID-19 patients, COVID-19 nucleic acid cycle threshold (CT value) of 56 patients≤30, while 31 patients >40.

**Fig. 2:**
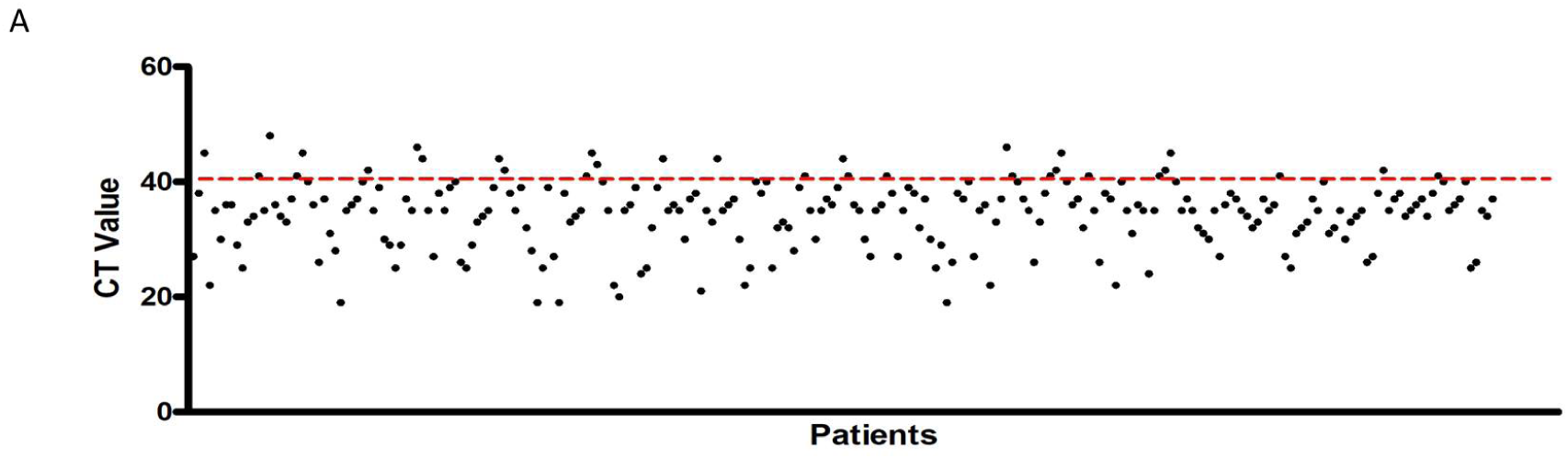
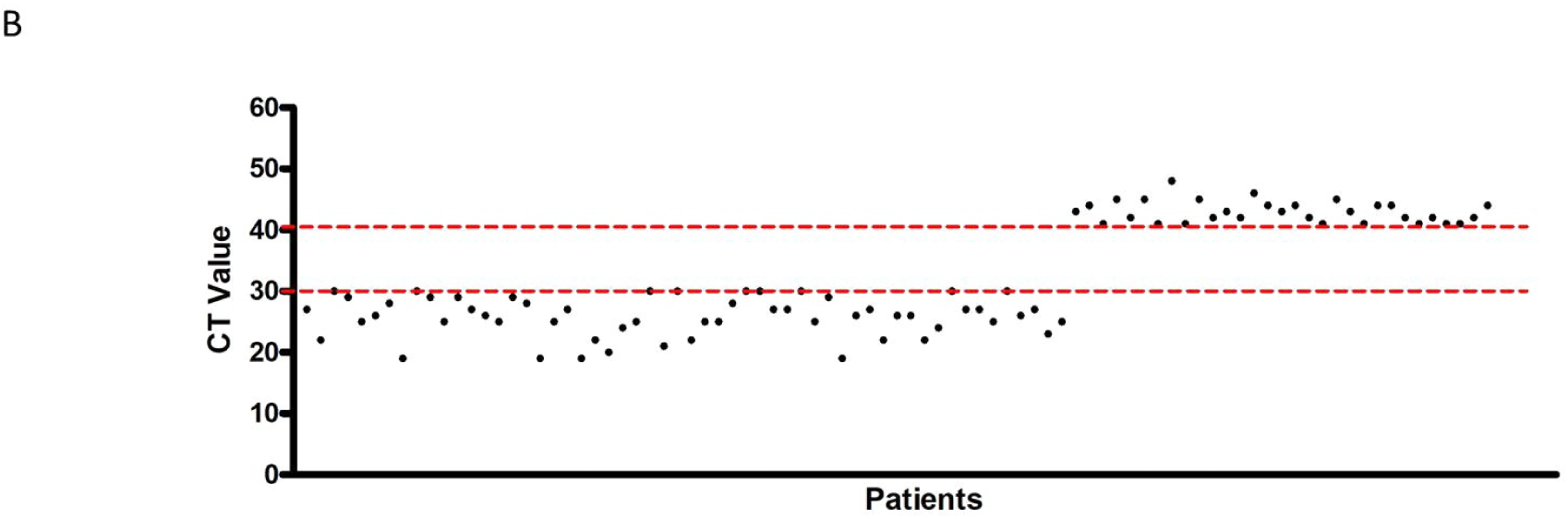
Proportion of participants with nucleic acid detection at different cutoff value. **(A)** Among 239 suspected COVID-19 participants, nucleic acid cycle threshold (CT value) of 208 patients≤40, while 31 participants >40. **(B)** Nucleic acid cycle threshold (CT value) of 56 patients ≤30, while 31 participants >40.

### 2. Nucleocapsid protein detection

We performed nucleocapsid protein (N antigen) detection of SARS-CoV-2 in nasopharyngeal swab samples in parallel with nucleic acid test in multiple centers. As shown in Figure 2, among the 208 patients with COVID-19 nucleic acid positive results, 141 were SARS-CoV-2 N antigen positive, with a sensitivity of 68%. Meanwhile, among the 31 patients with COVID-19 nucleic acid negative results, 31 were N antigen negative, with a specificity of 100%. In order to investigate the sensitivity and specificity of antigen detection methods from a clinical perspective, our study selected CT value ≤40 and ≤30 (Higher virus titer) as the nucleic acid positive group while CT value > 40 was used as the nucleic acid negative group. The results showed the positive coincidence rate (sensitivity) was 68%, the negative coincidence rate (specificity) was 100%,and the total coincidence rate (accuracy) was 72% when CT value≤40 as nucleic acid positive group while CT value > 40 as nucleic acid negative group (Fig3A). In addition, positive coincidence rate (sensitivity) was 98%, the negative coincidence rate (specificity) was 100%, and the total coincidence rate (accuracy) was 99% when CT value≤ 30 as nucleic acid positive group while CT value > 40 as nucleic acid negative group (Fig3B). It is of noteworthy that in this study cohort the earliest patient with fever in 3 days were identified by N antigen detection.

**Fig. 3:**
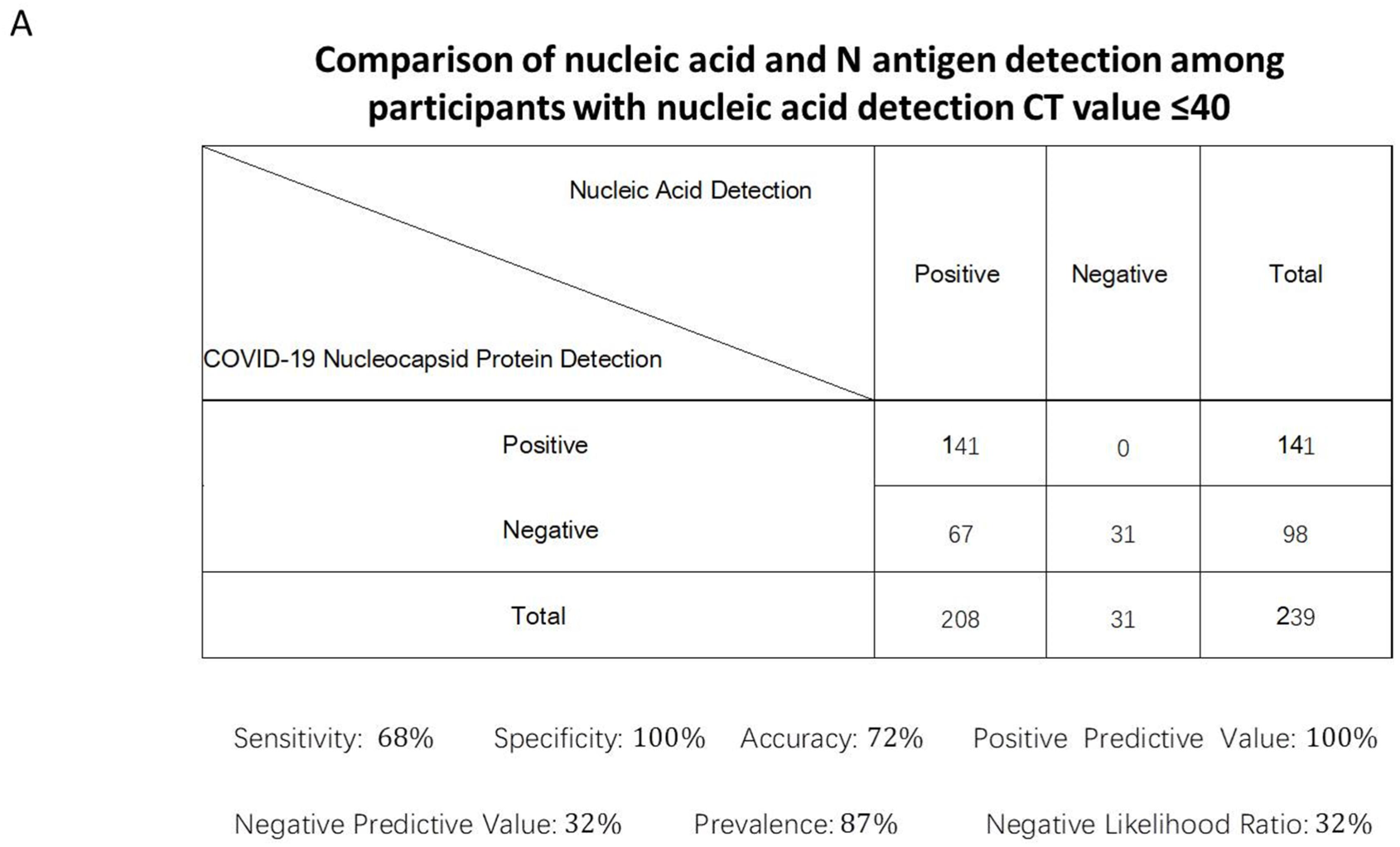
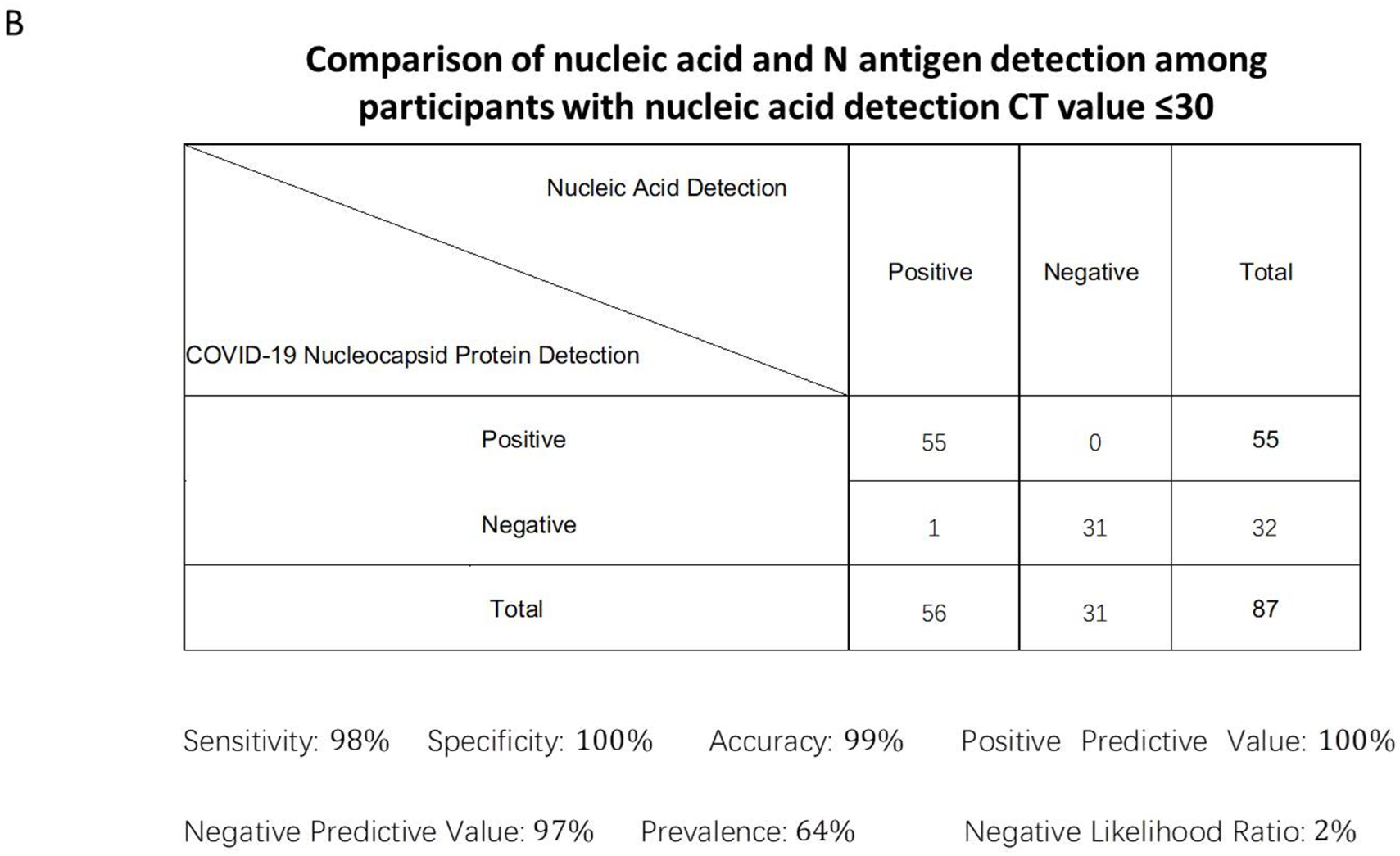
Comparison between nucleic acid detection and N antigen detection. **(A)** Comparison of nucleic acid and N antigen detection among participants with nucleic acid detection CT value ≤40. **(B)** Comparison of nucleic acid and N antigen detection among participants with nucleic acid detection CT value ≤30.

We performed N antigen detection of SARS-CoV-2 in urine in one in parallel with nucleic acid test of nasopharyngeal swab samples in same day. It is interesting that 14 of 19 (73.6%) participants with diagnosed COVID-19 were detected N antigen in urine (Table 1).

**Table 1:**
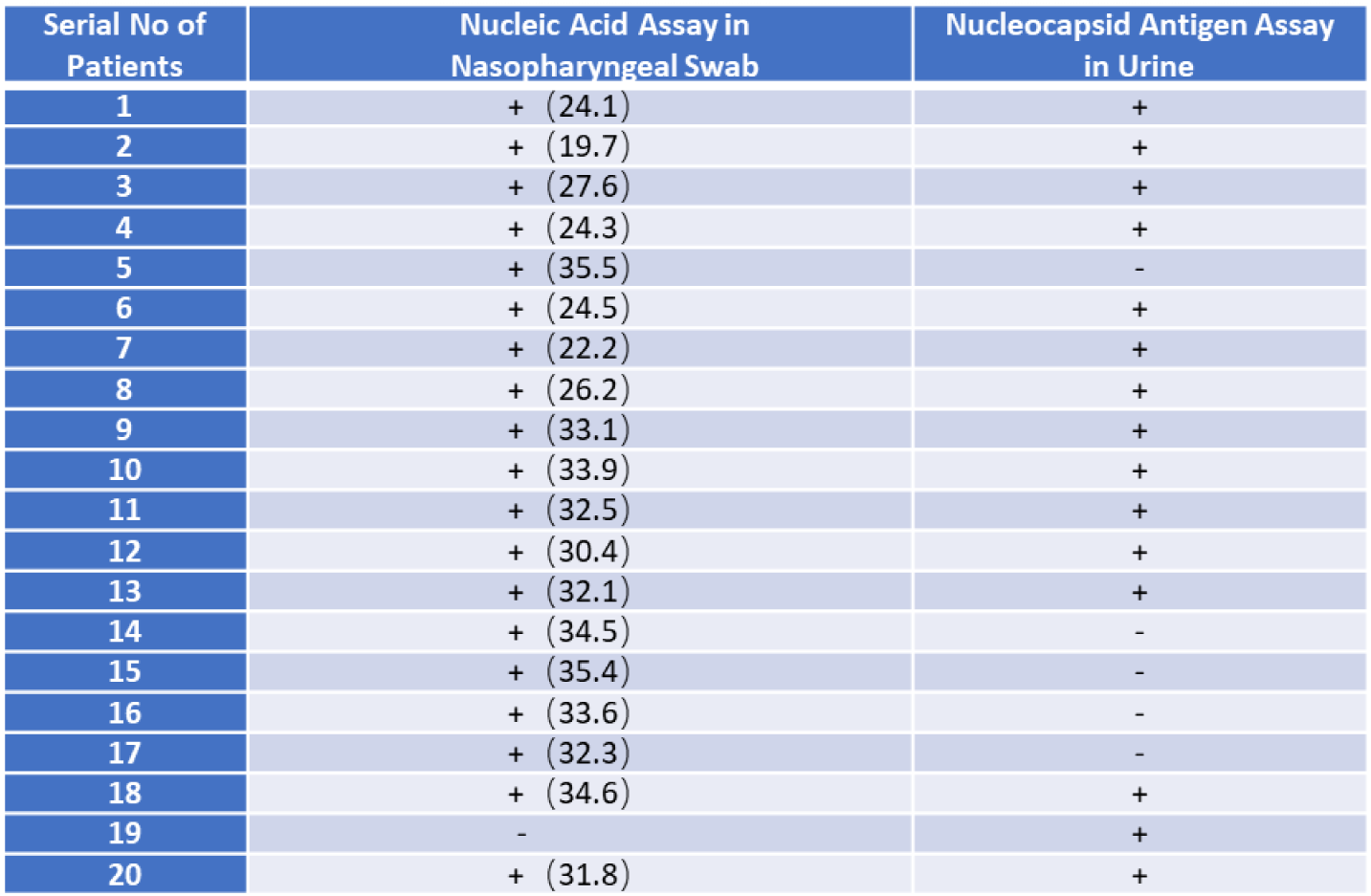
Examination of nucleocapsid antigen in urine compared by nucleic acid assay in nasopharyngeal swab. 14 0f 19(73.6%) the nucleic acid assay in nasopharyngeal swab positive patients exhibits nucleocapsid antigen positive in urine in same day. 1 nucleic acid assay positive patient exhibited nucleocapsid antigen positive in urine.

## Discussion

The current epidemic situation of COVID-19 still remains severe and it quickly spreads all over China and to more than 20 other countries. Early diagnosis is critical to prevent its spreading and clinical treatment. In practice, nucleic acid test and antibody assay have been employed in the diagnostic for suspected SARS-CoV-2 infections, but the use of novel coronavirus antigen for diagnosis has not been successfully developed. Now, the virus (SARS-CoV-2) nucleic acid RT-PCR test has become the standard method for diagnosis of SARS-CoV-2 infection due to its high detection rate [12]. However, nucleic acid test results are sometimes unstable and take too long, or can appear false negative and false positive. Moreover, IgM and IgG antibody detection method have been used in diagnosing with a relative higher positive rate [13]. However, virus-specific antibody usually produced in 2-4 weeks after infection.

Theoretically, viral antigen is the specific marker of the virus and precedes antibody appearance within the infected people. In our previous study in patients with severe acute respiratory syndrome (SARS), test of the N antigen of SARS-CoV achieved a sensitivity of 94% and 78% for the first 5 days and 6–10 days after onset, respectively. This pointed out that antigen detection has a high true positive rate and false negative rate, which can be used as an early diagnostic marker for SARS before 1 day clinical onset [11]. Other authors also indicate N antigen of SARS-CoV has a high sensitivity rate in early stage of SARS, which could be useful for early diagnosis of SARS [14], indicate antigen detection can play an important role in the early diagnosis. Therefore, detection of viral antigen can play a rapid screening effect and achieve the purpose of early diagnosis.

Here, we report a fluorescence immunochromatographic assay for detecting nucleocapsid protein of SARS-CoV-2 in nasopharyngeal swab samples and urine within 10 minutes, and evaluated it’s significance in diagnosis. In the double blind evaluation clinical trial with the nucleic acid test as the golden standard, we found that the nucleic acid test positive rate was 87% and 64%,depending on the CT value, the positive rate of N antigen detection of SARS-CoV-2 was 59 % and 63% separately, 100% of nucleocapsid protein positive and negative participants accord with nucleic acid test. However, N antigen detection achieved a sensitivity of 100%, which greatly reduced the false positive rate of nucleic acid detection. More importantly, the earliest patient after 3 days of fever can be identified by the method. Those findings indicate that our N antigen assay is an accurate, rapid, early and simple diagnosis method of COVID-19. It implies that this N antigen detection method not only guarantee early diagnosis in hospitals, and can also be used large-scale screening in community.

In an additional preliminary study, it is revealed that 73.6% of diagnosed patients with nucleic acid assay in nasopharyngeal swab detected nucleocapsid antigen in urine in same day. This coincides our finding of the SARS-CoV-2 invading kidney and test of N antigen in urine might be of diagnostic value [15].

## Data Availability

The data used to support the findings of this study are included within the article.

## Conflict of interest

The authors declare no financial or commercial conflict of interest.

## Authors’ contributions

Yuzhang Wu, Hongwei Zhou, Guohong Deng involved in the final development of the project and manuscript preparation; Yueping Liu, Zilin Yuan, Chao Han and Jiahui Chen analyzed the data; Bo Diao, Kun Wen and Jian Chen Jing Wang, Yunjie Dan, Yongwen Chen performed most of experiments; Yuxian Pan, Li Chen involved in the detection assay development.

## Ethics committee approval

This study was approved by the National Health Commission of China and Ethics Commission of General Hospital of Central Theatre Command and Hanyang Hospital.

## Notes

### Competing Interest Statement

The authors have declared no competing interest.

